# Uniform Manifold Approximation and Projection-based Assessment of Chronic Kidney Disease Aetiologies based on Urinary Peptidomics

**DOI:** 10.1101/2023.05.19.23290228

**Authors:** Emmanouil Mavrogeorgis, Tianlin He, Harald Mischak, Agnieszka Latosinska, Antonia Vlahou, Joost P. Schanstra, Lorenzo Catanese, Kerstin Amann, Tobias B. Huber, Joachim Beige, Harald Rupprecht, Justyna Siwy

**Affiliations:** Mosaiques Diagnostics GmbH, 30659 Hannover, Germany; Institute for Molecular Cardiovascular Research (IMCAR), RWTH Aachen University Hospital, 52074 Aachen, Germany; Center of Systems Biology, Biomedical Research Foundation of the Academy of Athens, 11527 Athens, Greece; Institut National de la Santé et de la Recherche Médicale (INSERM), U1297, Institute of Cardiovascular and Metabolic Disease, 31432 Toulouse, France; Université Toulouse III Paul-Sabatier, 31062 Toulouse, France; Department of Nephrology, Angiology and Rheumatology, Klinikum Bayreuth GmbH, 95447 Bayreuth, Germany; Kuratorium for Dialysis and Transplantation (KfH) Bayreuth, 95445 Bayreuth, Germany; Friedrich-Alexander-University Erlangen-Nürnberg, 91054 Erlangen, Germany; Department of Nephropathology, Institute of Pathology, Friedrich-Alexander-University of Erlangen-Nürnberg, 91054 Erlangen, Germany; III. Department of Medicine, University Medical Center Hamburg-Eppendorf, Hamburg, Germany; Hamburg Center for Kidney Health (HCKH), University Medical Center Hamburg-Eppendorf, Hamburg, Germany; Department of Infectious Diseases/Tropical Medicine, Nephrology/KfH Renal Unit and Rheumatology, St. Georg Hospital Leipzig, 04129 Leipzig, Germany; Kuratorium for Dialysis and Transplantation (KfH) Renal Unit, Hospital St. Georg, 04129 Leipzig, Germany; Department of Internal Medicine II, Martin-Luther-University Halle/Wittenberg, 06108 Halle (Saale), Germany

## Abstract

**Background:** The impact of artificial intelligence combined with advanced techniques is ever-increasing in the biomedical field appearing promising, among others, in chronic kidney disease (CKD) diagnosis. However, existing models are often single-aetiology specific. Proposed here is a pipeline for the development of single models able to distinguish and spatially visualize multiple CKD aetiologies.

**Methods:** Acquired were from the Human Urinary Proteome Database the urinary peptide data of 1850 healthy control (HC) and CKD (diabetic kidney disease-DKD, IgA nephropathy-IgAN, vasculitis) participants. The uniform manifold approximation and projection (UMAP) method was coupled to a support vector machine (SVM) algorithm. Binary (DKD, HC) and multiclass (DKD, HC, IgAN, vasculitis) classifications were performed, including or skipping the UMAP step. Last, the pipeline was compared to the current state-of-the-art single-aetiology CKD urinary models.

**Findings:** In an independent test set, the developed models (including the UMAP step) achieved 90.35% and 70.13% overall predictive accuracies, respectively, for the binary and the multiclass classifications (96.14% and 85.06%, skipping the UMAP step). Overall, the HC class was distinguished with the highest accuracy. The different classes displayed a tendency to form distinct clusters in the 3D-space based on their disease state.

**Interpretation:** Urinary peptide data appear to potentially be an effective basis for CKD aetiology differentiation. The UMAP step may provide a unique visualization advantage capturing the relevant molecular (patho)physiology. Further studies are warranted to validate the pipeline’s clinical potential in the presented as well as other CKD aetiologies or even other diseases.

## Introduction

The high prevalence and economic burden(1) of chronic kidney disease (CKD) underscore the vital need for further attempts on addressing its associated challenges. Failing to identify CKD in its former, asymptomatic stages, where therapy is expected to lead to improved outcome, eventually leads to an advanced disease state, in which an efficient aetiology-guided treatment is often obstructed by the phenotypic overlap between the various aetiologies. In that context, a CKD diagnosis is an essential and delicate issue. That said, a major clinical concern relates to the differential diagnosis of different CKD aetiologies, currently being suboptimal and in most cases relying on invasive kidney biopsies as the gold standard with, however, associated potential bleeding complications(2) and concerns about repetitiveness. Along these lines, several endeavors have been performed. For example, multiple binary classifiers had been employed for the discrimination of seven CKD aetiologies - including diabetic kidney disease (DKD) (mixed with nephrosclerosis), IgA nephropathy (IgAN) and vasculitis - using urinary peptidomics in a one-versus-all strategy(3). Even though this kind of studies demonstrate the value of machine learning in CKD diagnosis, aetiology-specific classifiers might lead to multiple positive hits, potentially resulting in an ambiguous diagnosis.

A major improvement of this strategy would be differential diagnosis by a single classifier specialized in multiple aetiologies. Nevertheless, among the most significant technical hurdles to overcome is the plethora of features detected in a relatively low number of observations, a situation well-known in the field as “curse of dimensionality”(4). This is exactly the case in CKD diagnosis, given the common elements of CKD aetiologies, the corresponding thousands of molecular determinants (e.g. peptides) detected as well as the often relatively low number (tens or hundreds at best) of analyzed samples.

The uniform manifold approximation and projection (UMAP)(5,6) is a recently developed non-linear dimensionality reduction method for the analysis of high-dimensional data that gains increasing popularity. The algorithm relies on two phases of analysis: first, a high-dimensional weighted graph of the data points is generated and subsequently its low-dimensional version is optimized. The UMAP algorithm provides a non-linear alternative to principal component analysis (PCA), although it lacks the PCA interpretability(5). Nevertheless, it provides several advantages over similar non-linear methods like t-SNE, such as superior run time performance(5,7) and more easily interpretable parameters. Last, UMAP shows a promising variety of applications in biological data interpretation, such as analyzing single-cell RNA sequencing and mass cytometry data(7).

This ability of machine learning algorithms to handle high-dimensional data combined with the resolving power of techniques that are able to identify plenty of molecular features during sample analysis could be highly relevant in CKD diagnosis. Along these lines, the analysis of urinary peptides and proteins based on capillary electrophoresis coupled to mass spectrometry (CE-MS) has been extensively applied for the identification and assessment of biomarkers in a number of diseases(8–11), relying on a completely non-invasive approach. The robustness of CE-MS has been highlighted in several studies(12–15). Applying urinary peptidomics combined with the support vector machine (SVM) algorithm demonstrated superior performance in comparison to the state-of-the-art(16). Several specific SVM-based peptide panels have been established in the field of chronic kidney disease (CKD), such as the IgAN237(17) or the CKD273(18) panels, the latter being recognized with a letter of support from the U.S. Food and Drug Administration (FDA)(19) and implemented in a clinical trial for early detection of diabetic kidney disease(20).

Building on available 1850 urine peptidomics datasets obtained from the Human Urinary Proteome Database(10), our aim was to establish a pipeline for the non-invasive differential diagnosis of CKD aetiologies in a novel approach, harnessing the dimensionality reduction and visualization capabilities of UMAP in a proof-of-concept study.

## Methods

Anonymized peptidomics data of 1850 urine samples corresponding to healthy controls (HC) and CKD participants of various aetiologies were acquired from the Human Urinary Proteome Database(10). The CKD aetiology selection criterion was based on a minimum of 70 samples. The HC samples were derived from participants without either signs of CKD or significant loss of kidney function (eGFR ≥ 60 mL/min/1.73m^2^) (n = 504). The CKD samples were derived from participants diagnosed with one of the following CKD aetiologies: IgAN (n = 737), DKD (n = 534), and vasculitis (n = 75). The study design is depicted in **Figure 1**. All datasets were from previously published studies and fully anonymized. The studies respected the regulations for protecting participants in medical research and the Declaration of Helsinki (2013). This study was approved by the ethics committee of the Friedrich-Alexander Universität Erlangen-Nürnberg, Germany (ethic approval code 264_20 B for the nephrological biobank and ethic approval code 221_20 B for the urinary proteomics analysis).

**Figure 1.**
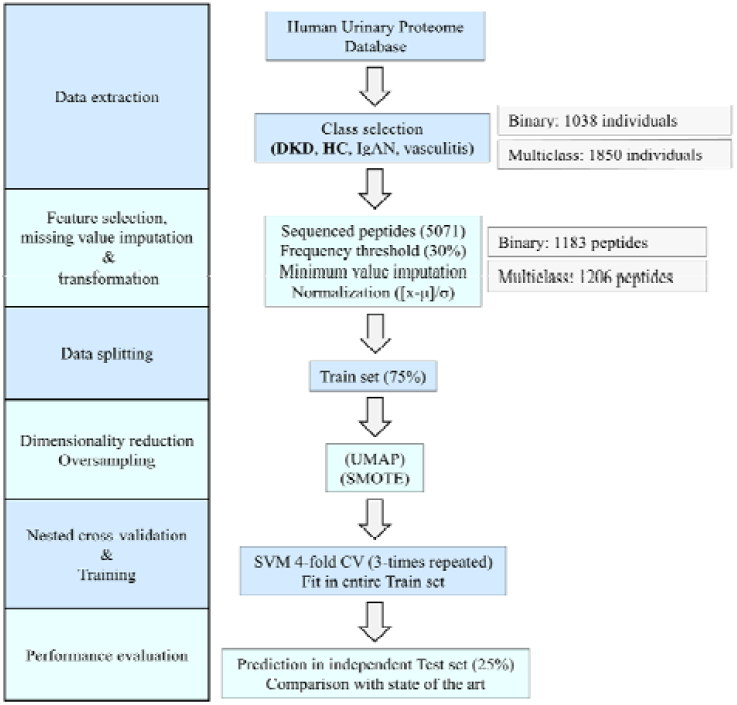
Study design. The urinary peptide datasets of a cohort of 1850 HC and CKD (DKD, IgAN, and vasculitis) individuals were implemented into a supervised machine learning pipeline for classification based on disease (or lack thereof). The pipeline was performed separately for DKD and HC classes (binary classification) as well as all classes (multiclass classification). Initially, a splitting of the dataset into a train (75%) and a test (25%) set was performed. Each time, the sequenced peptides present in at least 30% of the participants of the selected classes, were considered for further analysis and normalized ([x-mean(x)] / standard deviation(x)) after missing peptide values were imputed based on the respective minimum values, considering the train set only. A dimensionality reduction with the UMAP algorithm was performed (or skipped), while as an additional step during the training procedures in the multiclass classification only, the oversampling algorithm SMOTE(23) was applied. The later produced synthetic participants in all classes until a certain ratio of the (initially) majority class (i.e. IgAN) was achieved so as to account for the class imbalance. During a three-times repeated four-fold CV, SVM models were trained (in three out of four folds of the train set) and their performance was recorded (on the remaining fold) along the lines of an iterative search that relied on a Bayesian optimization(24) of the hyperparameters. The model that achieved the highest average accuracy across all the CV folds was selected as having the optimal combination of hyperparameter values. Subsequently, the selected model was trained in the entire train set and then tested for its predictive accuracy in the independent test set. μ: feature mean, σ: feature standard deviation, UMAP: Uniform Manifold Approximation and Projection, SMOTE: Synthetic Minority Over-sampling Technique. SVM: Support Vector Machines. CV: cross-validation in train set.

The methods used in this study are described in detail in the **appendix**. In brief, initially a urinary peptidomics protocol involving CE-MS, peptide sequencing and data evaluation was performed (**appendix p. 1**). UMAP(5,6) was applied to embed the data into a lower dimension (3D), as also described in several sources(21,22) and, for the multiclass classification only, oversampling(23) was used to account for the class imbalance (**appendix p. 2**). The entire peptidomics dataset was randomly split in train and test sets based on sample groups in a 75:25 ratio for classification purposes (**appendix pp. 2-3**). The train set features with the disease labels, were fed to a SVM classifier to generate a model able to distinguish participants based on their disease state. To this end, models were trained within a three-times repeated, four-fold cross validation (CV), in which Bayesian optimization(24) was used as an iterative search method in the context of hyperparameter tuning. The machine learning pipeline was based on R statistical software (**appendix p. 3**).

## Results

Urinary peptidomics data of 1850 individuals were extracted from the Human Urinary Proteome Database(10). This set included 504 HC participants as well as 534 patients with DKD, 737 with IgAN, and 75 with vasculitis for which the available clinical information is presented in **Table 1**. A machine learning pipeline was implemented to discriminate DKD and HC (binary classification) as well as DKD, HC, IgAN, and vasculitis (multiclass classification) with the main goal to maximize the separation of the participants based on their CKD aetiology diagnosis (class). The binary differentiation was used as an initial evaluation to explore the UMAP capabilities before moving on to the more complex challenge of multiple CKD aetiologies differentiation. In this first step, DKD was selected, because it is the most common CKD aetiology; in addition, in our dataset the DKD and HC classes were balanced in terms of sample sizes. The study design is illustrated in **Figure 1**.

**Table 1.**
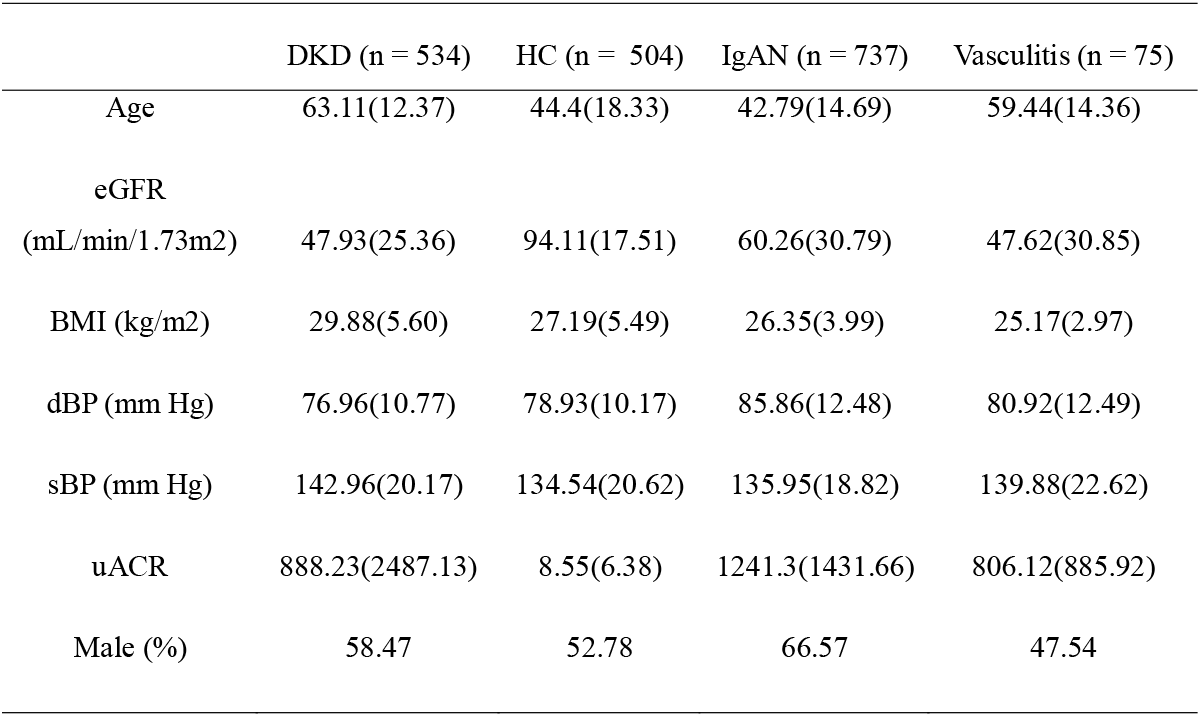
Cohort clinical characteristics. Given is the number of the participants of the entire classes. For the clinical characteristics each time a mean (standard deviation) or percentage is displayed, as calculated based on the available participant clinical information. M: Male. eGFR: estimated glomerular filtration rate. uACR: urinary albumin to creatinine ratio. BMI: body mass index. dBP: diastolic blood pressure. sBP: systolic blood pressure.

|  | DKD (n = 534) | HC (n = 504) | IgAN (n = 737) | Vasculitis (n = 75) |
| --- | --- | --- | --- | --- |
| Age | 63.11(12.37) | 44.4(18.33) | 42.79(14.69) | 59.44(14.36) |
| eGFR<br>(mL/min/1.73m<sup>2</sup>) | 47.93(25.36) | 94.11(17.51) | 60.26(30.79) | 47.62(30.85) |
| BMI (kg/m<sup>2</sup>) | 29.88(5.60) | 27.19(5.49) | 26.35(3.99) | 25.17(2.97) |
| dBP (mm Hg) | 76.96(10.77) | 78.93(10.17) | 85.86(12.48) | 80.92(12.49) |
| sBP (mm Hg) | 142.96(20.17) | 134.54(20.62) | 135.95(18.82) | 139.88(22.62) |
| uACR | 888.23(2487.13) | 8.55(6.38) | 1241.3(1431.66) | 806.12(885.92) |
| Male (%) | 58.47 | 52.78 | 66.57 | 47.54 |

### Binary classification: differentiation of DKD and HC classes

Initially, UMAP was applied as a standard unsupervised dimensionality reduction method to the peptidomics data of 534 DKD and 504 HC participants in an attempt to visualize their potential separation in the 3D-space **(Figure 2A)**. Although the majority of the participants of the same class diagnosis appeared to be clustered together, a substantial overlap of the clusters did not allow for a clear separation. That observation indicated, on one hand, the utility of UMAP in embedding high-dimensional urinary peptidomics data in a low-dimensional space, and on the other that a supervised UMAP approach may be more relevant for class separation. Thereby, the UMAP algorithm was applied with the diagnosis label (supervised UMAP), which led to a major class separation improvement **(Figure 2B)**. In a next step, the default supervised UMAP hyperparameters (specifically k-neighbors and minimum distance), were also tuned along the SVM hyperparameters during a three times-repeated four-fold CV procedure using the train set and the model with the combination of hyperparameter values with the best performance was, finally, selected. The selected model achieved 89.89% average accuracy across all the folds during the train set CV, while in the independent test set, an overall 90.35% predictive accuracy was achieved, the latter being excluded from the training procedures, and thus representing an unbiased source of the model’s efficiency. The UMAP embeddings of the train and test sets are illustrated in **Figure 2C-D**. The per-class accuracies of the model for both the train set CV and the independent test set are illustrated in **Figure 2E**.

**Figure 2.**
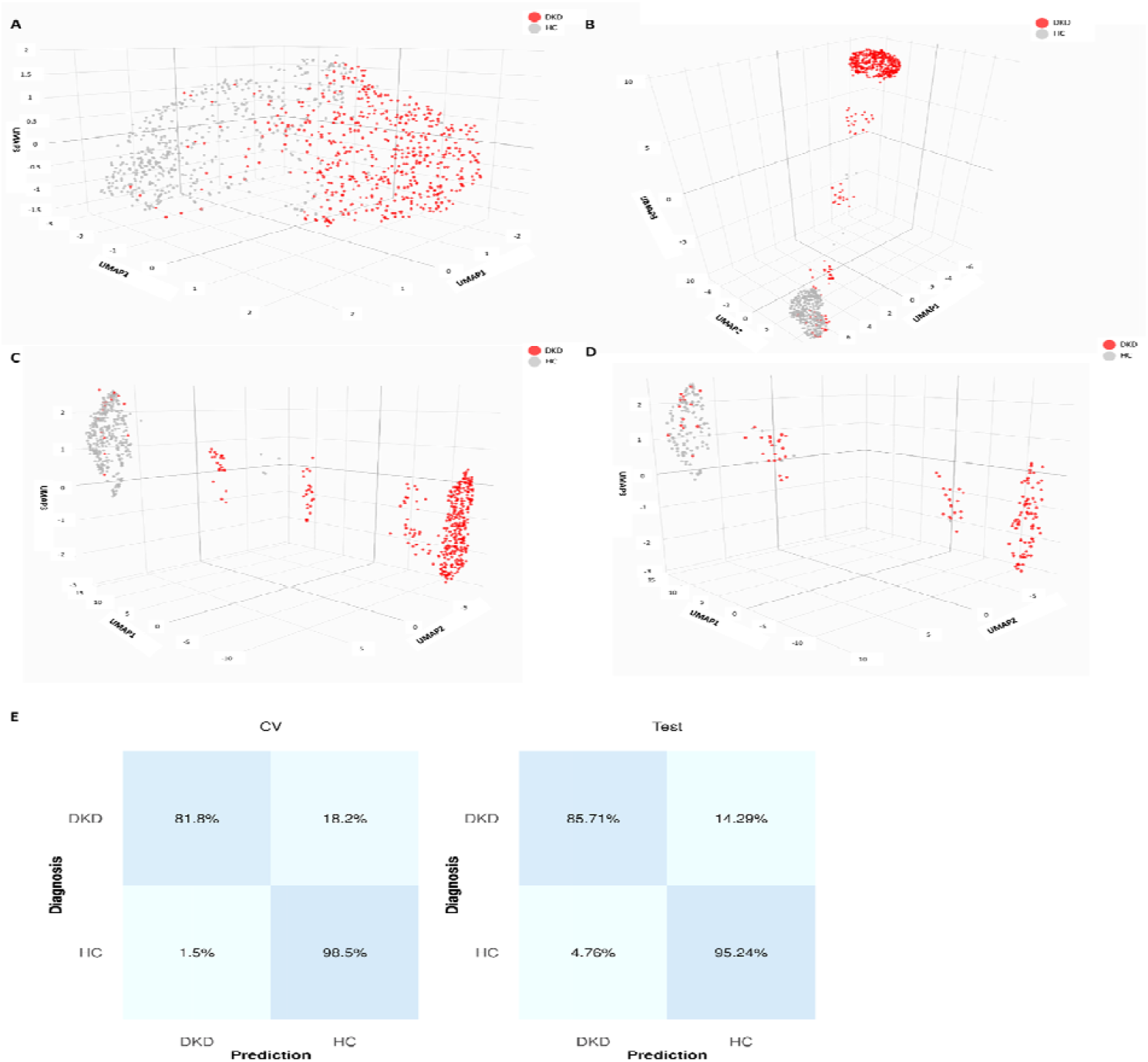
Binary classification results. The whole peptidomics profiles of DKD (red) and HC (gray) participants in the 3D-space were used as a basis for the default parameters of the UMAP algorithm in its (A) unsupervised as well as (B) supervised version. Cluster formation was more evident when the supervised UMAP with tuned parameters was performed, as observed when applied on the (C) train set and (D) independent test set embeddings. E) Confusion matrices based on the results of the train set cross-validation (CV, average across all folds) as well as the predictions in the independent test set. Classification accuracies are displayed in percentages.

### Multiclass classification: differentiation of multiple CKD aetiologies and HC classes

Subsequently, the same pipeline was utilized to differentiate all four classes: DKD, HC, IgAN, and vasculitis. Again, UMAP (default parameters) was applied to the data, which highlighted a tendency of cluster formation, although without a clear separation **(Figure 3A)**. This was to a substantial degree improved in the respective supervised UMAP embeddings **(Figure 3B-D)**. To adjust for the numeric imbalance of these classes, an oversampling approach was implemented during the training procedures. The overall performance of the selected model across all the CV folds of the train set (average of 74.18%) as well as the predictions in the independent test set (70.13%), were recorded. In detail, predictions in the independent test set displayed accuracies of 56.39%, 66.30%, and 78.95% for DKD, IgAN, and vasculitis classes, respectively, achieving the highest accuracy (88.89%) in differentiating the HC class from CKD aetiologies. **(Figure 3E)**.

**Figure 3.**
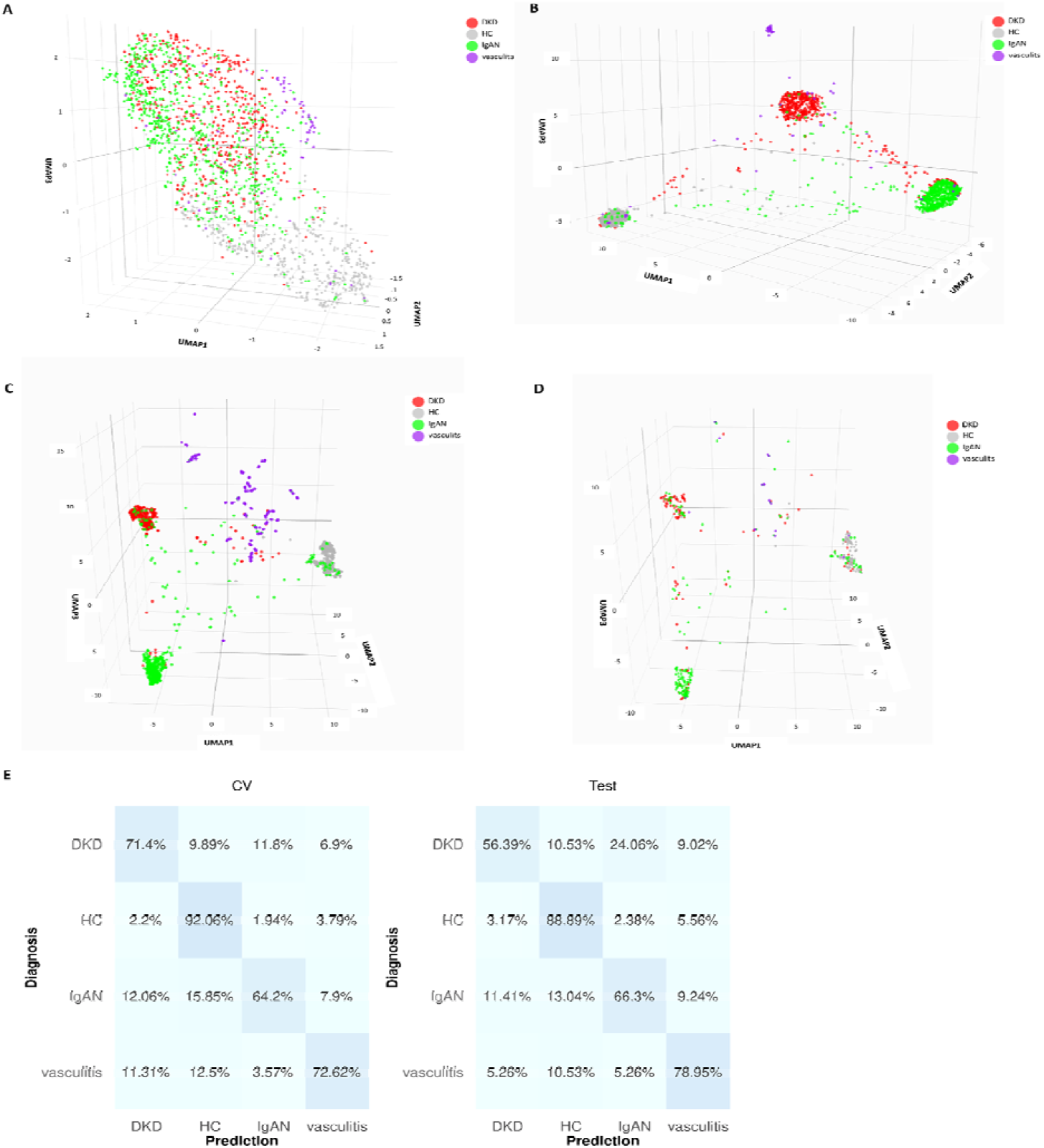
Multiclass classification results. The whole peptidomics profiles of DKD (red), HC (gray), IgAN (green), and vasculitis (purple) participants in the 3D-space were used as a basis for the UMAP algorithm (default parameters) in its (A) unsupervised as well as (B) supervised version. Cluster formation was more evident when the supervised UMAP with tuned parameters was performed, as observed by the (C) train set and (D) independent test set embeddings. (E) Confusion matrices based on the results of the train set cross-validation (CV, average across all folds) as well as the predictions in the independent test set. Classification accuracies are displayed in percentages. Of note, an oversampling step was performed during the training procedures.

### Comparison with SVM-only classifier

In an attempt to illustrate the added value of UMAP as an important dimensionality reduction step in urinary peptidomics as well as the proposed pipeline as a whole, additional comparisons were performed. Initially, a SVM model was built and trained as described above, but skipping the UMAP step. In the binary classification, the selected model displayed an overall accuracy of ≥ 95.56% in both the train set CV (average across all folds) and the independent test set (**Figure 4A**). In the multiclass classification, the model achieved during the train set CV an overall average accuracy of 87.51%, while the overall prediction accuracy in the independent test was 85.06% and the per-class accuracies were 86.47%, 82.61%, and 63.16% for DKD, IgAN, and vasculitis, respectively (**Figure 4B**). Of note, in the latter classification, the HC class was distinguished with 90.48% accuracy.

**Figure 4.**
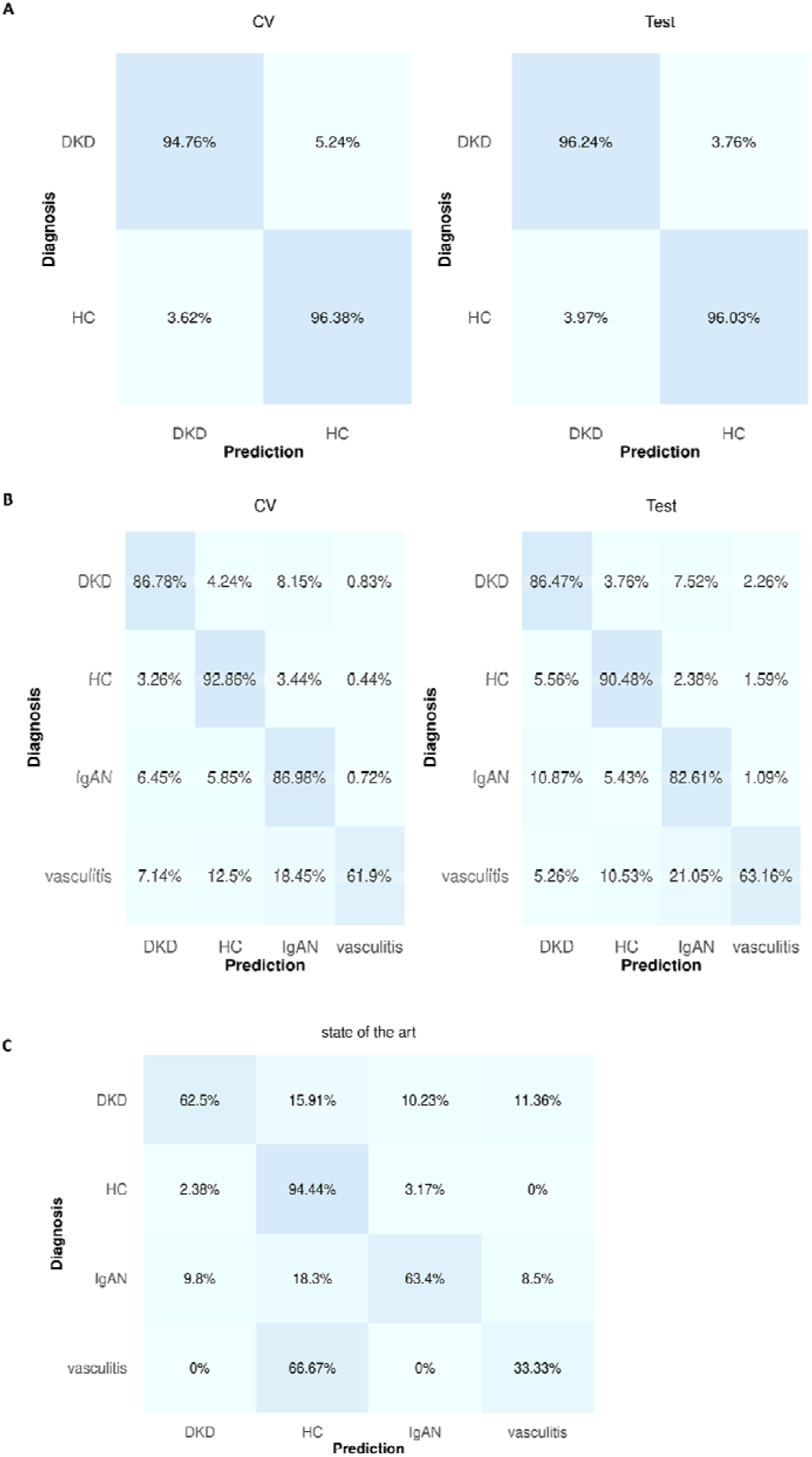
Comparison without including UMAP in the pipeline as well as with the current state of the art. Confusion matrices of the predictions in (A) binary and (B) multiclass classifications. (C) Predictions using the current state-of-the-art single-aetiology classifiers(3,18).

### Comparison with the state of the art in CKD urinary proteomics

Subsequently, the comparison with the individual CKD-aetiology models described in Siwy et al. (2017)(3) was performed. Considered were the classifiers specific for DKD (mixed with nephrosclerosis), IgAN, and vasculitis classes, since these aetiologies were relevant in the current study. Predictions were made only for the 373 participants of the independent test set (n = 462) that had not been a part of the train set of the CKD differential diagnosis classifiers in Siwy et al. (2017)(3); specifically these corresponded to: 88 DKD, 126 HC, 153 IgAN, and 6 vasculitis individuals. To differentiate HC from CKD patients, the CKD273(18) classifier was utilized. The models correctly predicted 62.50%, 94.44%, 63.40%, and 33.33% of the DKD, HC, IgAN, and vasculitis classes, respectively (**Figure 4C**).

## Discussion

The invasive kidney biopsy is still the gold standard in the (differential) diagnosis and therapeutic decisions in CKD, despite its limitations. Since biopsy is an invasive procedure with potential post-surgery complications, inadequate sampling, disagreement in the interpretation between pathologists, dependence on appropriate organ size, and impracticalities in repetitiveness, the implementation of specific biomarkers appears highly relevant. To this end, several efforts have been performed to identify biomarkers that could non-invasively support the CKD differential diagnosis. Along these lines, the existing literature was explored using the terms: “chronic kidney disease”, “CKD”, “kidney disease”, “differential diagnosis”, “types”, “aetiolog*”, “etiolog*”, “classifier*”, and “panel”. That said, the research of CKD differential diagnosis seems to be mainly focused on genetic studies aiming at sequencing, such as CKD-related genetic panels(25–33). These studies at times confirmed the presence of suspected inherited kidney diseases and even led to a correction of the traditional diagnosis. Nevertheless, the presented diagnostic performance varied in parameters, such as the onset (congenital to adult), CKD stage (early to end) and specific kidney disease type. Along these lines, considering that ∼10% of adult CKD is attributed to hereditary origin, a meticulous record of the patient clinical information (e.g. phenotype, family history) before the genetic test was recommended. Last, potential technique constraints (e.g. variants within promoter regions are undetected by whole exome sequencing) may also have to be considered when assessing the correspondent diagnostic yield of genetic tests, although in part these can be addressed, such as in the ongoing whole genome sequencing study by Soraru et al. (2022)(34).

On the other hand, relevant attempts on the proteomics/peptidomics field, often relying on machine learning approaches, have been scarcely performed. Glazyrin and colleagues (2020)(35), demonstrated that using urine samples, nephrosclerosis was distinguished from the mixed DKD and glomerulonephritis classes, with the latter two being subsequently differentiated from each other using plasma samples. Although displayed high classification performance, the study was based on only 34 participants, and thus among others, a separation in train and test sets was not feasible. This was implemented by Fernando et al. (2019)(36), nevertheless, focusing only in differentiating CKD of unknown aetiology from a mixed CKD aetiology class (DKD, nephrosclerosis, glomerular diseases). In the largest proteomics/peptidomics study based on our search, Siwy et al. (2017)(3) using a cohort of total 1180 participants, developed several single-aetiology models for seven CKD aetiologies (plus one control class for which the CKD273(18) was used), embodying the current state-of-the-art. The classifier performance achieved a 0.77 area under the curve or higher in the independent test set. Nevertheless, although a classifier specialized on a single aetiology, could in an appropriate scientific design, potentially, demonstrate substantial specialization towards that aetiology, contradictory positive hits as produced by multiple single-aetiology classifiers have a rather ambiguous diagnosis contribution. Thus, a single classifier for distinguishing multiple aetiologies appears highly clinically relevant.

In the current work, we demonstrated that supervised UMAP coupled with the SVM algorithm can be utilized as a tool to differentiate multiple CKD aetiologies based solely on urinary peptidomics data. In both the binary (DKD, HC) and the multiclass (DKD, HC, IgAN, vasculitis) classifications, there was a tendency of distinct cluster formation in the 3D-space appearing to be representative of the diagnosis state. The standard deviation (sd) of the selected metric that was recorded during the three-times repeated four-fold CV in both classifications was fairly small **(Supplementary table 1)**, indicating that the performance of the pipeline was independent of the different fold combinations. The model performance was each time also confirmed in an independent (from the training procedures) test set.

The overall model performance in the binary classification was superior to the respective in the multiclass one. This was expected, since the binary task was less complex, basically distinguishing CKD of one aetiology (DKD) from HC. That said, the HC class was distinguished with the highest success in both the binary and multiclass classifications. This can be attributed to the fact that HC participants are pathologically more distant from the CKD classes, which was also spatially observed in the UMAP plots (**Figure 3 and 4**), thus justifiably being distinguishable from the rest of the classes. This observation can be interpreted as further evidence for the validity of the presented approach. Considering the multiclass classification, the initial class imbalance in the train set along the small vasculitis size and the selected metric (accuracy, due to its simplicity), indicated that the application of an oversampling approach could be appropriate for a pathophysiology-centered model instead of one potentially favoring the majority class. In that particular classification (including the UMAP step), during predictions on the independent test set, the selected model had the lowest performance distinguishing DKD, assigning a substantial part of its participants to the former majority class, IgAN. Nevertheless, although DKD and IgAN are not that clinically similar, their routine treatment involves several common aspects, especially the anti-hypertension treatment involving angiotensin-converting-enzyme inhibitors and angiotensin II receptor blockers, as well as the recently implemented, SGLT-2 inhibitors.

As a dimensionality reduction method, UMAP is able to remove, at least to a degree, the noise of a dataset and thus increase the model performance. Considering the SVM-alone pipeline, this was not observed here, which we speculate may be linked to the noise reduction already achieved via applying the 30% peptide frequency threshold considering only the sequenced peptides as part of the routine analysis in the peptidomics field. In that way, from tens of thousands of peptides consistently being detected in urine, only 1183 and 1206 were considered for further analysis in the binary and multiclass classifications, respectively. In that context, using UMAP to further reduce the feature space (and thus, the correspondent dataset information) to only 3 spatial coordinate features resulted, not entirely unexpectedly, in a model of poorer predictive performance on the independent test set (∼70% accuracy) (as well as in terms of train set CV mean and sd) in comparison to the SVM-alone approach (∼85% accuracy); but still appeared to be able to retain a substantial part of the dataset’s “information”. Consequently, in different scientific designs in which an efficient feature selection/removal method is not available, the inclusion of UMAP in the pipeline could potentially contribute to noise removal and thus, to higher model performance. That said, the dimensionality reduction along with its spatial, single-sample, visualization properties constitute UMAP a substantial step in such pipelines. Of note, in the binary classification, UMAP demonstrated on the independent test set a predictive performance (∼90%) close to the respective of the SVM-alone approach (∼96%).

Considering that proteomic/peptidomic studies are scarce in CKD differential diagnosis, the presented pipeline was compared to the DKD (mixed with nephrosclerosis), IgAN, and vasculitis classifiers of the aforementioned study(3), using the CKD273 for the HC group(18). As expected from the anticipated difference in molecular pathology, the HC class was the most highly distinguishable. The presented pipeline (including the UMAP step) demonstrated, in comparison to these single-aetiology classifiers, fairly comparable performance, outperforming the vasculitis class, although the latter may as well be attributed to luck, since only six vasculitis patients were tested in this single-aetiology classifier. Other differences in performance can also be, among others, attributed to the train sets not being identical, different pipelines (e.g. selecting features based on differential expression(3) instead of 30% frequency threshold, alternative hyperparameter optimization etc.) as well as the initial number of features (sequenced peptides) during model development, as per availability in each case.

The presented study has limitations, some of which may be improved in future work. First, class balance was not the case for the multiclass classification. Class imbalance is hardly avoidable when working with such retrospective datasets, among others due to the inherent difference in disease prevalence. Therefore, as noted, an oversampling approach was used in an attempt to enhance the pathophysiology-centered nature of the classifier by compensating for inappropriate effects (against the minority classes) as a potential result of the class imbalance. Further, the here investigated CKD aetiologies represent only a fraction of the broad CKD spectrum, and thus the inclusion of additional aetiologies in a larger study seems well justified. It is also expected that the inclusion of relevant clinical parameters may increase model performance. However, due to incomplete clinical records of some participants, this could not be implemented. Last, only the SVM (radial basis kernel) classifier was assessed. As a result, using larger cohorts with aetiologies across the disease spectrum, ideally of equal class size and aetiology-representative, in combination with further/alternate preprocessing steps, more extensive hyperparameter optimization, classifier and metrics assessment as well as also considering clinical parameters, could potentially enable improved model performance.

Nevertheless, the current study highlighted at the same time several interesting clinical aspects. While omitting the UMAP step led to higher classification performance, a major advantage of UMAP as a dimensionality reduction method (additionally to the potential noise removal) is the cohort visualization in low-dimensional space irrespective of the initial number of features (e.g. peptides). This is especially practical for high-dimensional omics data, in which thousands of features per sample are detected. To our knowledge, this work is the first of its kind to reduce the complex urinary peptidome to such a degree that participants of multiple CKD aetiologies are efficiently presented as single data points in space, collectively forming distinct diagnosis clusters. As such, this visualization tool in an appropriate pipeline could complementarily contribute to the context of clinical assessment of CKD. It is even more tempting to speculate that the position in UMAP is based on the specific individual molecular determinants, which may potentially be highly valuable in determining personalized intervention by predicting drug response, considering also several relevant clinical parameters, such as clinical characteristics, medical history and disease confounders, progression, microbiome, diet, exercise etc. This hypothesis will be investigated in more detail and would ultimately have to be proven in an appropriately-powered randomized clinical trial. Last, the presented pipeline appears to be able to generate models capable of distinguishing different CKD aetiologies and thus, might complementarily aid or even, ideally, rather point at times towards a potential avoidance of the kidney biopsy.

In conclusion, in this proof-of-concept study, we established a robust and relatively fast pipeline for simultaneous classification of multiple CKD aetiologies and visualization of individual samples in the 3D-space based on urinary peptides. This could be specifically relevant when invasive procedures are typically avoided (in early stage disease, at high risk, for repeat sampling) or not possible at all.

Given the inherent risks, shortcomings and invasive character of kidney biopsy, determining the CKD aetiology in a non-invasive way appears highly relevant. The presented differentiation and visualization approach can be used as a supplementary tool in clinical practice, not only for the presented, but likely also for additional CKD aetiologies. In that way, it could complementarily aid diagnosis or even, ideally, potentially lead towards avoidance of the kidney biopsy. Along these lines, it is to be expected that this approach, by the inclusion of additional datasets of high-dimensional omics data combined with artificial intelligence, will see routine use in modern health care systems. Last, its potential clinical utility, possibly supported by including additional clinical parameters, may pave the way for a more robust, non-invasive, disease monitoring or response to treatment prediction, thus supporting therapeutic decisions.

## Data Availability

Data sharing
Data will be made available upon request directed to the corresponding author. Proposals will be reviewed and approved by the investigators and collaborators based on scientific merit. After approval of a proposal, data will be shared through a secure online platform after signing the data access and confidentiality agreement.
Code Availability
Code was generated based on the functions in the respective R packages as described in the methods and will be made available upon request directed to the corresponding author.

## Author contributions

HM, EM, JS, JB, HR and TH conceptualized the study. EM and TH designed the study methodology. EM, TH, AL and JS performed the investigation and EM and JS performed the validation. HR and JB had access to and verified the data reported in this study. EM and TH wrote the first draft of the manuscript, visualization was done by EM. The supervision was performed by HR, HM, JB, JS, AV and JPS. All authors revised successive drafts of the manuscript, and approved the final version.

## Funding

This work was supported in part by the European Union’s Horizon 2020 research and innovation programs (860329 Marie-Curie ITN “STRATEGY-CKD” as well as 764474 Marie-Curie ITN “CaReSyAn”). The German Research Foundation also supported in part this work (SFB/TRR219 Consortium Project ID: 322900939). This work was also supported by BMBF founded project UPTAKE (01EK2105A-E). AV, JS, HM and JPS are members of the COST (European Cooperation in Science and Technology) action PERMEDIK CA21165.

All underlying studies were conducted to conform to regulations on the protection of individuals participating in medical research and accordance with the principles of the Declaration of Helsinki and had received ethical approval from the responsible institutional review boards. Written informed consent was obtained from all participants at the time of sampling. All data sets received were anonymized.

## Data sharing

Data will be made available upon request directed to the corresponding author. Proposals will be reviewed and approved by the investigators and collaborators based on scientific merit. After approval of a proposal, data will be shared through a secure online platform after signing the data access and confidentiality agreement.

## Code Availability

Code was generated based on the functions in the respective R packages as described in the methods and will be made available upon request directed to the corresponding author.

