## Appendix 1 for "Uniform Manifold Approximation and Projection-based Assessment of Chronic Kidney Disease Aetiologies based on Urinary Peptidomics"

Supplement to: *Uniform Manifold Approximation and Projection-based Assessment of Chronic Kidney Disease Aetiologies based on Urinary Peptidomics*

Emmanouil Mavrogeorgis, Tianlin He, Harald Mischak, Agnieszka Latosinska, Antonia Vlahou, Joost P. Schanstra, Lorenzo Catanese, Kerstin Amann, Tobias B. Huber, Joachim Beige, Harald Rupprecht, Justyna Siwy

**Table of contents**

**Urinary proteomics……………………1**

**Dimensionality reduction……………...2**

**Oversampling…………………………..2**

**Classification…………….……………..2**

**Statistical software………..……………3**

**References………..………..………..…..3**

**Supplementary table 1………..………..5**

**Urinary peptidomics**

The urine samples have been prepared and measured by CE-MS as stated before(1–4). The P/ACE MDQ capillary electrophoresis system (Beckman Coulter, USA) connected to a micro-TOF-MS (Bruker Daltonics, Germany) was used for the peptide analysis. The probabilistic clustering algorithm along with isotopic distribution and conjugated masses for charge has been used for raw MS data evaluation as described previously(1). Twenty-nine fragments of collagens that are generally not affected by disease were used for the normalization of the CE-MS signal intensity(5). To obtain the amino acid sequence, urinary peptides were fragmented using Orbitrap MS coupled to CE (CE-MS/MS) or liquid chromatography (LC–MS/MS)(6). The fragmentation spectra were matched to the protein sequences from up-to-date databases (International Protein Index, Reference sequence database at NCBI and UniProt Knowledgebase) using Proteome Discoverer 1.4 (Thermo Scientific, Bremen, Germany) with an integrated Sequest search engine). The following search parameters were applied: (1) precursor mass tolerance: 5 ppm, (2) fragment mass tolerance: 50 mDa and (3) variable post-translational modifications: hydroxylation of proline and oxidation of methionine, and (4) no specific cleavage. Before classification, the peptide signal intensities of the selected classes were normalized based on the formula “[x-mean(x)] / standard deviation(x)” after the missing values were imputed based on the respective minimum values calculated in the train set. Only the sequenced peptides detected in at least 30% of the participants were considered for further analysis, namely, 1183 and 1206 peptides for the binary and multiclass classifications, respectively.

**Dimensionality reduction**

Uniform Manifold Approximation and Projection (UMAP)(7,8) was applied to embed the data into a lower dimension (3D), also explained very well in several sources(9,10). In brief, UMAP is a non-linear dimensionality reduction method that relies in two phases. Initially, a high-dimensional weighted graph of the data is generated. Each data point is characterized by an outer radius, the width of which is not fixed, but is based on its distance from its k nearest neighbors. In that way, data points can form a connection when their radii overlap, but the algorithm decreases the likelihood of connection (represented by edge weights), the larger a radius grows. In that way, data points connected with highly weighted edges are more likely to remain close together. Subsequently, a low-dimensional version as structurally proximal as possible to the former high-dimensional graph is optimized. Two of the most important parameters in UMAP are the k number of neighbors and the minimum distance. Considering larger values in the former parameter favors capturing the “big picture” of the data structure, while a lower value, favors capturing the finer details of it. Minimum distance refers to the proximity of the data points in the low-dimensional space with lower values tending to lead in more densely packed data points. The k-nearest neighbors and minimum distance parameters of UMAP were tuned based on the SVM classification performance as described below.

**Oversampling**

The Synthetic Minority Over-sampling Technique (SMOTE)(11) was utilized to produce synthetic participants based on k neighbors, until each class reached a certain ratio in relation to the initial number of the majority class (i.e. IgAN) in the train set. Of note, due to oversampling, the whole train set (n = 1388) ended up consisting of 2648 participants of equally-sized classes (n = 662). Oversampling did not affect the independent test set, which consisted of 462 participants: 133 DKD, 126 HC, 184 IgAN, 19 vasculitis participants. No oversampling was performed in the binary classification due to its balanced classes and thus, the initial train (n = 779) and test (n = 259) sets remained intact.

**Classification**

The entire dataset was randomly split in train and test sets in a 75:25 ratio. The train set features with the disease labels, were fed to a SVM classifier to generate a model able to distinguish participants based on their disease state. In detail, using several combinations of hyperparameter values generated through an iterative grid search method, namely, Bayesian optimization(12), the potential models were trained in three folds of the train set with their performance being recorded after class predictions on the fourth fold. When this was performed in total three times (with different fold separation in train set) for all the potential models and the different sets of fold combinations, the average (across all folds) performance was calculated per model. The model with the highest average accuracy was selected as the optimal one. Accuracy appeared as the appropriate metric in this proof-of-concept study due to its simplicity given also the equally-sized classes in the binary as well as the multiclass (the latter as a result of the oversampling) classifications. The hyperparameters that were considered for tuning were the following: the k-nearest neighbors and minimum distance (for UMAP(7,8) as well as cost and sigma (for SVM). In the multiclass classification, SMOTE(11) served as an oversampling method to compensate for class imbalance and its hyperparameters k-neighbors and oversampling ratio were also tuned. The hyperparameter values as well as model performances during train set CV and predictions are gathered in **Supplementary table 1**. In case multiple models during train set CV demonstrated the same average accuracy, the ones with the lowest standard deviation found in the latest iteration were selected as optimal. Exploring the utilization of several potential aspects in the current pipeline (e.g. pre-processing steps or even reproducibility indexes etc.), the ones that resulted in the highest train set CV performance in the multiclass classification (including the UMAP step) were applied for all classifications. Of note, a combined grouped-stratified resampling was not the case for the vasculitis class potentially given its low size and/or sample groups and thus, the vasculitis patient percentage varied across the CV folds, before applying the oversampling.

**Statistical software**

The results and findings of the current paper were based on R programming (R version 4.2.2, R Foundation for Statistical Computing, Vienna, Austria) running on Ubuntu 22.04 computer software. The R script also heavily relied on the collection of R packages in the tidyverse(13) package, e.g. plotting the confusion matrices. In that context, the machine learning pipeline was built based on the tidymodels(14) package, the capabilities of which, are described in detail elsewhere(15). The data were split in train and test sets using the function group_initial_split() and the train set was used for repeated cross validation based on the function group_vfold_cv(), filling in both functions the arguments *strata* and *group* with the diagnosis and patient identification number columns, respectively. For the multiclass classification the argument *pool* = 0 was added in the aforementioned functions given the low size of the vasculitis class. The workflow was performed using as an engine the kernlab package(16) for the SVM classifier and as a preprocessing step the UMAP(7,8) algorithm of the embed(17) package. Last, as an oversampling algorithm, SMOTE(11) was utilized based on the package themis(18). The Bayesian optimization(12) was performed with the function tune_bayes() of the dials(19) package with parameters: initial = 20, iter = 80 and no_improve = 80. The UMAP plots were based on the plotly(20) package. The confusion matrices were arranged based on the ggpubr package(21). The results can be reproduced in R by setting the function set.seed (2020) before the relevant functions.

**Supplementary table 1**

Hyperparameter values along with respective model performances during the train set CV (average across all folds) as well as with regards to the independent test set predictions, for both the binary and multiclass classifications. Inside the parentheses it is indicated whether a hyperparameter characterizes the SMOTE, UMAP or SVM. Iter refers to the respective iteration during the Bayesian optimization of iterative search.

SMOTE: smote synthetic minority over-sampling technique. UMAP: uniform manifold approximation and projection. SVM: support vector machines. CV: cross validation.
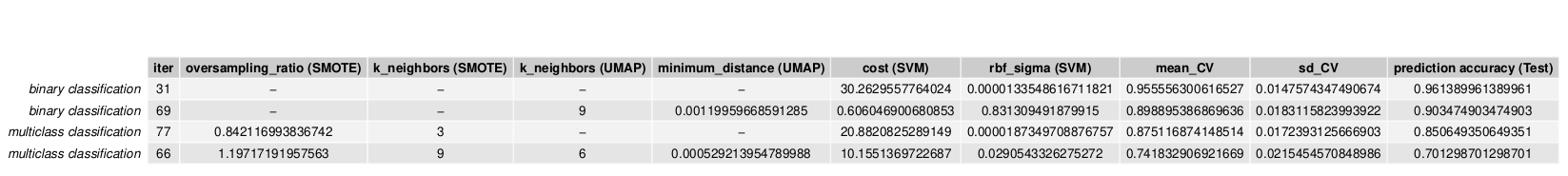
